# Epidemiology of stillbirths and maternal post-discharge outcomes following stillbirths in Uganda: a prospective cohort study

**DOI:** 10.64898/2026.01.23.26344736

**Authors:** Obed Twinamatsiko, Vuong Nguyen, Clare Komugisha, Angella Namala, Joseph Ngonzi, Nathan Kenya Mugisha, Yashodani Pillay, Astrid Christofferson-Deb, Matthew O Wiens

**Author notes:** Corresponding authors: Matthew O. Wiens. Institute for Global Health, 305 – 4088 Cambie Street, Vancouver, BC, V5Z 2X8, Canada.

## Abstract

**Background:** Stillbirth remains a critical public health challenge in low-resource settings and is a significant cause of perinatal mortality, where gaps in antenatal care, healthcare access, and socioeconomic disparities exacerbate risks. Understanding maternal post-discharge outcomes and identifying modifiable predictors for stillbirths is essential to improve care pathways.

**Methods:** This prospective cohort study analyzed maternal and perinatal health data from 7131 women who delivered 7359 newborns at the Mbarara and Jinja Regional Referral Hospitals between April, 2022 and September, 2023. A stillbirth was defined as the death of a foetus >28 weeks of gestation. Univariate logistic regression models were used to determine risk factors.

Odds ratios (ORs) with 95% confidence intervals (CIs) were reported.

**Results:** Among 7129 women who survived and were discharged post-delivery, 261 (3.7%) experienced a stillbirth. Six-week post-discharge readmission of women of stillborn infants was 8.5% as compared to 3% among those who were discharged with a live newborn. The strongest risk factor for a stillbirth was previous child death (OR: 7.11, 95% CI: 5.5–9.17, *p* <0.001), followed by transport delays >1 hour (OR: 2.71, 95% CI: 1.95–3.75) and pregnancy-related illnesses (OR: 1.70, 95% CI: 1.30–2.25). Each additional year of maternal age increased odds by 2% (OR: 1.02, 95% CI: 1.00–1.04). Protective factors included adequate antenatal care (4–8 visits) (OR: 0.51, 95% CI: 0.40–0.66) and partner support (OR: 0.72, 95% CI: 0.56–0.95).

**Conclusion:** Maternal morbidity following stillbirths is high. Furthermore, several demographic, health, and socioeconomic strongly influenced the risk of stillbirths. Many stillbirths may be prevented following early identification of these risk factors through interventions to ensure expectant mothers receive adequate support during their pregnancy.

## Introduction

Stillbirth remains a profound health challenge in low-resource settings, exacting a heavy toll on families and healthcare systems. Sub-Saharan Africa bears the highest proportion of the global stillbirths, accounting for over two-thirds of the world’s estimated 2 million annual stillbirths, despite some efforts made since 2000 (1,2).

Research in low– and middle-income countries (LMIC), primarily conducted through facility-based studies and population-based surveillance systems, health information systems, and systematic reviews, has identified several key factors that predict stillbirth, including advanced maternal age, higher parity, inadequate antenatal care, and socioeconomic disadvantage (3,4). Previous research at Mulago Hospital in Uganda identified specific predictors of intrapartum stillbirths, such as previous pregnancy loss, referral delays, and delivery complications, but these findings are limited to labor-related events and lack insight into antepartum stillbirths occurring earlier in pregnancy (5). Despite these findings, significant gaps persist. Many studies rely on retrospective or cross-sectional designs with limited granularity on modifiable factors like transport barriers, social support, or specific health conditions, which vary widely across contexts (6). Moreover, distinctions between fresh and macerated stillbirths, indicating timing and potential etiology, are rarely explored in large cohorts, leaving uncertainties about differential risk profiles that could guide targeted interventions.

While these and other studies have advanced our understanding of factors associated with stillbirth, post-discharge trajectories among women experiencing a stillbirth remain understudied, with existing literature focusing primarily on mental health impacts such as depression, anxiety, and grief. Clinical outcomes, such as recurrent illness or mortality, are insufficiently described from an epidemiologic perspective. This gap limits our ability to address the full spectrum of needs among women following a stillbirth in resource-limited settings.

This study leverages a prospective cohort of women delivering at two Ugandan regional referral hospitals to address these challenges. Originally designed to examine post-discharge outcomes in mother-newborn dyads, this secondary analysis aims to identify candidate predictors of stillbirths while also providing insight into the post-discharge trajectories of mothers who have experienced a stillbirth.

## Methods

### Study Design Setting

This study is a secondary analysis of a prospective study in which participants were recruited from April 01, 2022, to September 30 2023, which included a six-week post-discharge follow-up period (7). Mothers and newborns were enrolled from two regional referral facilities in Uganda: Mbarara Regional Referral Hospital in Southwestern Uganda and Jinja Regional Referral Hospital in Eastern Uganda. Together, these facilities cover two different districts in Uganda and serve a population of approximately 973,000 people, providing an acceptable representation of the population outside of the capital city of Kampala.

#### Ethics approval

This study was approved by Makerere University School of Public Health (MakSPH) Research and Ethics Committee (SPH-2021-177), the Uganda National Council of Science and Technology (UNCST) in Uganda (HS2174ES), and the University of British Columbia in Canada (H21-03709). This study has been registered at clinicaltrials.gov (NCT05730387). This manuscript is reported using the Strengthening the Reporting of Observational Studies in Epidemiology (STROBE) guidelines (8).

#### Participants

Eligibility was defined as any woman or adolescent girl over 12 who lived in the catchment districts of the two facilities and was admitted for delivery. Ineligible individuals included those who lived in refugee settlements, could not provide their consent, were not admitted for delivery, had a language barrier, or lacked access to a phone. Written informed consent was obtained from all study participants, including minors, whose consent was provided by their guardians or legal parents. A quasi-random sample technique was used because census sampling was not practical, enabling the enrollment of seven to ten individuals per site each day.

#### Data Collection

Data collection tools and study procedures have been previously described and are available through the Smart Discharges Study Dataverse (9). Briefly, all participants received routine care at the discretion of their medical team. In cases where study nurses noted concerning signs or symptoms, these were reported to the medical team. Following delivery and obtaining informed written consent, trained study nurses collected data grouped according to four periods of care; admission, delivery, discharge, and six-week post-discharge follow-up. Data from admission and delivery was captured from the hospital medical record where possible and by direct observation, direct measurement or patient interview when not. To describe socioeconomic status, we used the Global Network Socioeconomic Status Index score from 0-10 developed using the multidimensional poverty index (MPI) and subsequently validated in previous studies (10–12). The newborn status (dead or alive) was recorded at birth, with stillbirths defined as the death of a foetus in utero after >28 weeks of gestation based on the definitions provided by the WHO. Stillbirths were further subdivided into fresh or macerated stillbirths, subjectively defined by the research nurse.

Data were collected using a custom-built application installed on Android tablets with an encrypted database and uploaded daily to a Research Electronic Data Capture (REDCap) database hosted at the BC Children’s Hospital Research Institute in Vancouver, Canada.

#### Statistical Analysis

All analyses were conducted using R Statistical Software (R version 4.4.1) and RStudio version (RStudio, Boston, MA, USA) (https://posit.co/products/open-source/rstudio/). Descriptive statistics were generated to summarize baseline characteristics and clinical, social, or maternal variables, stratified by stillbirth status (yes/no) and further stratified by fresh vs. macerated stillbirth. Continuous variables were reported as means with standard deviations or medians with interquartile ranges (IQR), while categorical variables were expressed as counts and percentages. Univariable logistic regression models were used to assess the association between potential risk factors (e.g., maternal age, antenatal visits, transport times) and stillbirth outcomes. For fresh and macerated stillbirth comparisons, a separate logistic model was fitted to identify risk factors specific to each stillbirth subtype. Effect sizes were presented as odds ratios (OR) with 95% confidence intervals (CI). Statistical significance was defined as p < 0.05.

Missing data were handled via complete-case analysis unless otherwise specified. All results are provided in tabular form to highlight key demographic, clinical, and outcome variables across stillbirth groups.

## Results

Of the 9,497 pregnant women screened for eligibility, 7,131 were enrolled, resulting in 7,359 births. Overall, 2 (0.03%) maternal deaths occurred during delivery, and 7,129 women were discharged: 7,060 (99.0%) routinely, 32 (0.4%) against medical advice, 33(0.5%) discharged for other reasons, and 4 (<0.1%) had missing discharge status (Figure 1). A total of 261 (3.7%) of mothers who were discharged had a stillbirth (3.8% of total births). Among these, 133 (51%) of mothers had fresh, and 114 (43.7%) had macerated stillbirths, with 14 unclassified. The stillbirth rate was 38 per 1,000 births.

**Fig 1:**
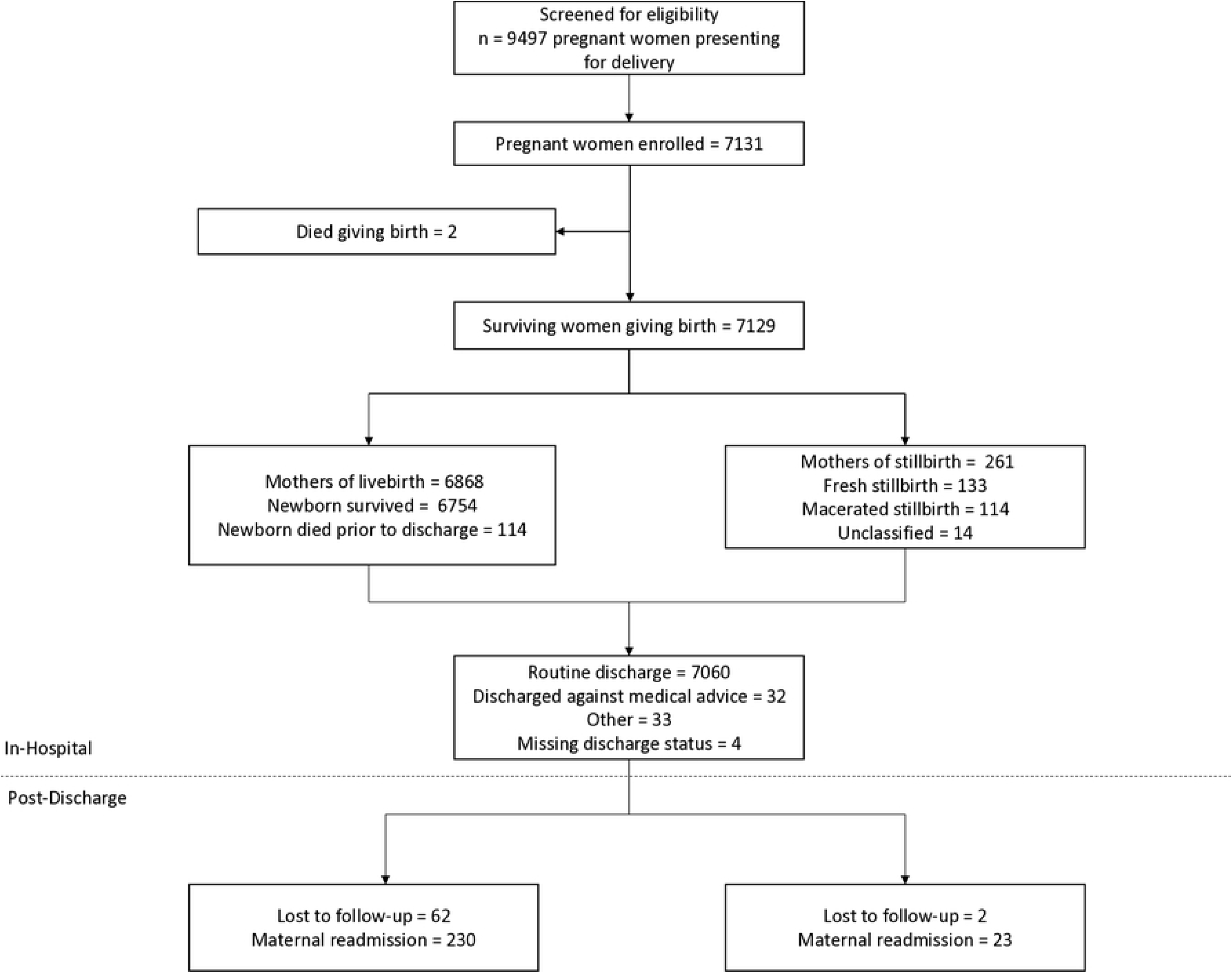
Study enrollment flow diagram. Flow diagram of participant enrollment, pregnancy outcomes, discharge status, and post-discharge follow-up. Discharge status includes all surviving mothers regardless of pregnancy outcome (live birth or stillbirth).

### Post-discharge risk for women following stillbirth or in-hospital newborn death

Women who experienced a stillbirth showed the highest cumulative rate of post-discharge maternal readmission, reaching 8.5% within 42 days after discharge (Figure 2). Women who delivered liveborn infants who died before discharge had a similarly high risk at 6%, while women whose babies survived to discharge had the lowest risk, at 3%.

**Fig 2:**
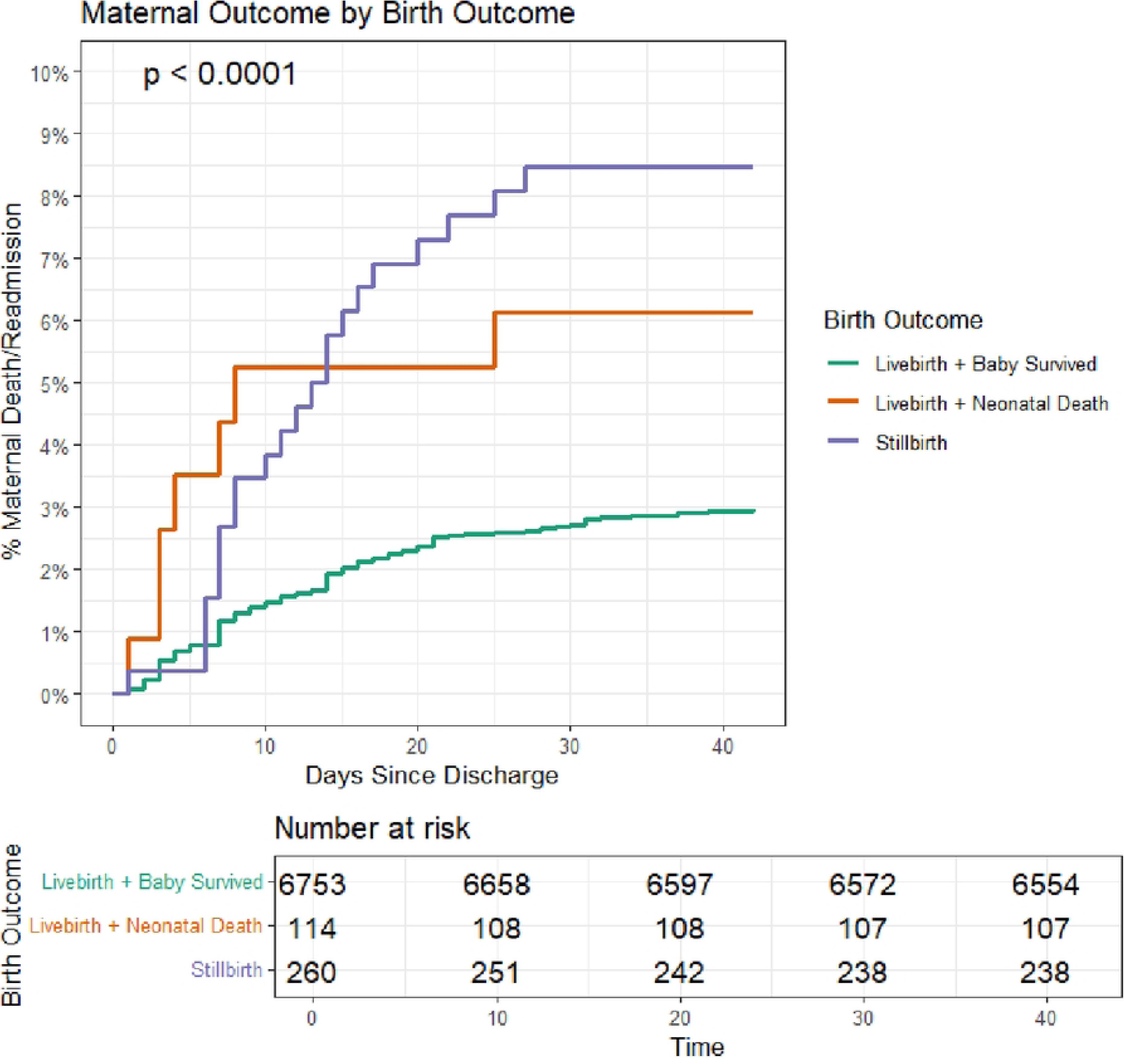
Cumulative incidence survival curve by birth outcome and neonatal outcome. Kaplan–Meier cumulative incidence curves comparing the post-discharge risk of adverse outcomes among mothers who had a live birth, stillbirth, or in-hospital neonatal death. Curves display the time-to-event distribution over the follow-up period.

### Factors associated with stillbirth

A full list of variables and their associations with stillbirth is summarized in Table 1. Pregnancy factors associated with stillbirth included maternal age (OR: 1.02, 95% CI: 1–1.04, p = 0.035), parity (OR: 1.16, 95% CI: (1.08–1.24, p < 0.001), and having a diagnosis of chronic illness (OR: 1.58, 95% CI: 1.13–2.17, p = 0.006) or pregnancy-related illness (OR: 1.70, 95% CI: (1.30–2.25, p < 0.001). Delivery related features such as preterm delivery (delivery 28 – <37 weeks) (OR: 4.01, 95% CI: 2.97–5.34, p < 0.001), meconium in amniotic fluid (OR: 2.12, 95% CI: 1.18–3.52, p = 0.006), and more than one baby delivered (OR: 1.96, 95% CI: 1.1–3.25, p = 0.014) were also associated with stillbirths.

**Table 1:** Demographic, social, clinical, and delivery-related characteristics of mothers with stillbirths and live births, and univariate odds ratios for stillbirth. The data are presented as odds ratios (OR) with 95% confidence intervals (CI), estimated using a univariate logistic regression model with live birth as the reference outcome. An OR >1 indicates higher odds of stillbirth. Continuous variables are expressed as mean (SD) or median (Q1, Q3), while categorical variables are shown as n (%). The reference category for categorical variables is labeled as “Ref.” For non-binary categorical variables, the global p-value appears in the reference row. Missing data percentages are provided for each variable.

| Variable | Stillbirth | Livebirth | n (%)<br>Missing | Odds Ratio (95% CI) | P-value |
| --- | --- | --- | --- | --- | --- |
| **N Mothers** | 261 | 6868 |  |  |  |
| **Demographic and Social** |  |  |  |  |  |
| **Maternal age, mean (SD) years** | 27.3 (6.4) | 26.5 (5.9) | 2 (0.03%) | 1.02 (1, 1.04) | 0.035 |
| **Lives with the father of the baby** | 223 (85.4%) | 6215 (90.5%) | 5 (0.07%) | 0.61 (0.44, 0.88) | 0.006 |
| **Education level** |  |  | 5 (0.07%) |  |  |
| *≤P3* | 15 (5.7%) | 248 (3.6%) |  | Ref. | <0.001 |
| *P4-P7* | 105 (40.2%) | 2157 (31.4%) |  | 0.80 (0.48, 1.46) | 0.445 |
| *S1-S6* | 119 (45.6%) | 3080 (44.8%) |  | 0.64 (0.38, 1.15) | 0.112 |
| *Post-secondary* | 22 (8.4%) | 1378 (20.1%) |  | 0.26 (0.14, 0.53) | <0.001 |
| **Food insecurity during pregnancy** | 54 (20.7%) | 1188 (17.3%) | 5 (0.07%) | 1.25 (0.91, 1.68) | 0.159 |
| **Have any children who have died** | 120 (46%) | 734 (10.7%) | 4 (0.06%) | 7.11 (5.5, 9.17) | <0.001 |
| **Socioeconomic Index Score, mean (SD)** | 2 (1, 4) | 3 (2, 4) | 4 (0.06%) | 0.81 (0.75, 0.88) | <0.001 |
| **Transport time** |  |  | 3 (0.04%) |  |  |
| *Less than 30 minutes* | 88 (33.7%) | 3325 (48.4%) |  | Ref. | <0.001 |
| *30 minutes to 1 hour* | 106 (40.6%) | 2606 (37.9%) |  | 1.54 (1.15, 2.05) | 0.003 |
| *More than 1 hour* | 67 (25.7%) | 934 (13.6%) |  | 2.71 (1.95, 3.75) |  |
| **Transport mode** |  |  | 3 (0.04%) |  |  |
| *Motorized transport* | 87 (33.3%) | 2088 (30.4%) |  | Ref. | <0.001 |
| *Non-motorized transport* | 135 (51.7%) | 4370 (63.6%) |  | 0.74 (0.56, 0.98) | 0.033 |
| *Ambulance* | 39 (14.9%) | 407 (5.9%) |  | 2.30 (1.54, 3.38) | <0.001 |
| **Delayed <1 hour coming to the hospital** |  |  |  |  |  |
| *No delay* | 207 (79.3%) | 5976 (87%) | 0 (0%) | 0.57 (0.42, 0.79) | <0.001 |
| *Terrain* | 7 (2.7%) | 42 (0.6%) | 0 (0%) | 4.48 (1.82, 9.45) | <0.001 |
| *Transport Cost* | 23 (8.8%) | 517 (7.5%) | 0 (0%) | 1.19 (0.75, 1.8) | 0.442 |
| *General transport delay* | 19 (7.3%) | 245 (3.6%) | 0 (0%) | 2.12 (1.27, 3.35) | 0.002 |
| *Weather(rain)* | 1 (0.4%) | 19 (0.3%) | 0 (0%) | 1.39 (0.08, 6.72) | 0.751 |
| *Other* | 10 (3.8%) | 129 (1.9%) | 0 (0%) | 2.08 (1.01, 3.81) | 0.028 |
| **Pregnancy and Health** |  |  |  |  |  |
| **Parity, median (Q1, Q3)** | 2 (1, 4) | 2 (1, 3) | 5 (0.07%) | 1.16 (1.08, 1.24) | <0.001 |
| **Pregnancy loss per child, median (Q1, Q3)** | 0 (0, 0) | 0 (0, 0) | 5 (0.07%) | 1.13 (0.94, 1.33) | 0.179 |
| **Diagnosed with chronic illness before pregnancy** | 46 (17.6%) | 819 (11.9%) | 0 (0%) | 1.58 (1.13, 2.17) | 0.006 |
| **Diagnosed with pregnancy-related illness during pregnancy** | 187 (71.6%) | 4103 (59.7%) | 0 (0%) | 1.70 (1.3, 2.25) | <0.001 |
| **Previous caesarean** | 63 (24.1%) | 1556 (22.7%) | 4 (0.06%) | 1.09 (0.81, 1.44) | 0.578 |
| **Diagnosed with HIV** |  |  | 0 (0%) |  |  |
| *No HIV* | 248 (95%) | 6429 (93.6%) |  | Ref. | 0.479 |
| *HIV Before Pregnancy* | 12 (4.6%) | 375 (5.5%) |  | 0.83 (0.44, 1.43) | 0.534 |
| *HIV During Pregnancy* | 1 (0.4%) | 64 (0.9%) |  | 0.41 (0.02, 1.84) | 0.371 |
| **Referral** | 189 (72.4%) | 2973 (43.3%) | 3 (0.04%) | 3.44 (2.62, 4.55) | <0.001 |
| **Admitted to hospital during pregnancy** | 46 (17.6%) | 955 (13.9%) | 4 (0.06%) | 1.32 (0.95, 1.82) | 0.091 |
| **Number of antenatal visits attended** |  |  | 4 (0.06%) |  |  |
| <4 | 104 (39.8%) | 1757 (25.6%) |  | Ref. | <0.001 |
| 4-8 | 151 (57.9%) | 4988 (72.6%) |  | 0.51 (0.4, 0.66) | <0.001 |
| >8 | 6 (2.3%) | 119 (1.7%) |  | 0.85 (0.33, 1.82) | 0.709 |
| **Delivery Maternal** |  |  |  |  |  |
| **Caesarean delivery** | 175 (67%) | 4386 (63.9%) | 3 (0.04%) | 0.87 (0.67, 1.13) | 0.297 |
| **Delivery at <37 weeks** | 66 (25.3%) | 535 (7.8%) | 0 (0%) | 4.01 (2.97, 5.34) | <0.001 |
| **Meconium in amniotic fluid** | 15 (5.7%) | 194 (2.8%) | 18 (0.25%) | 2.12 (1.18, 3.52) | 0.006 |
| **Vaginal Exams** |  |  | 22 (0.31%) |  |  |
| 0-2 | 126 (48.3%) | 3572 (52%) |  | Ref. | 0.233 |
| 3-5 | 109 (41.8%) | 2517 (36.6%) |  | 1.23 (0.94, 1.59) | 0.124 |
| >5 | 25 (9.6%) | 758 (11%) |  | 0.94 (0.59, 1.42) | 0.763 |
| PROM | 25 (9.6%) | 283 (4.1%) | 187 (2.62%) | 2.54 (1.61, 3.83) | <0.001 |
| **More than one baby delivered** | 15 (5.7%) | 207 (3%) | 2 (0.03%) | 1.96 (1.1, 3.25) | 0.014 |
| **Discharge Maternal** |  |  |  |  |  |
| **Admitted to higher level of care for post-delivery complications** | 19 (7.3%) | 549 (8%) | 4 (0.06%) | 0.90 (0.54, 1.41) | 0.674 |
| **Discharge systolic blood pressure**, mean (SD) mmHg | 114.3 (13) | 114.6 (11.8) | 72 (1.01%) | 1.00 (0.99, 1.01) | 0.672 |
| **Discharge diastolic blood pressure**, mean (SD) mmHg | 75.8 (10) | 75.6 (9.5) | 72 (1.01%) | 1.00 (0.99, 1.01) | 0.763 |
| **Respiratory rate**, mean (SD) breaths per minute | 19.5 (3.5) | 19.4 (3.5) | 91 (1.28%) | 1.01 (0.97, 1.05) | 0.568 |
| **Temporal artery temperature**, mean (SD) °C | 36.3 (0.4) | 36.3 (0.4) | 79 (1.11%) | 1.36 (0.99, 1.87) | 0.059 |
| **Best SpO2**, mean (SD) % | 98.6 (1.2) | 98.5 (1.3) | 84 (1.18%) | 1.10 (0.99, 1.24) | 0.085 |
| **Best heart rate**, mean (SD) beats per minute | 96.2 (15.5) | 94.5 (15.3) | 78 (1.09%) | 1.01 (1, 1.02) | 0.080 |
| **Hematocrit**, mean (SD) % | 35.4 (5.1) | 37.7 (4.7) | 93 (1.3%) | 0.92 (0.89, 0.94) | <0.001 |
| **Discharge destination, own home** | 175 (67%) | 5215 (75.9%) | 0 (0%) | 0.64 (0.5, 0.84) | 0.001 |
| **Support** | 79 (30.3%) | 1494 (21.8%) | 0 (0%) |  |  |
| *Mother* |  |  |  | 1.56 (1.19, 2.04) | 0.001 |
| *Mother-in-law* | 21 (8%) | 644 (9.4%) | 0 (0%) | 0.85 (0.52, 1.3) | 0.469 |
| *Husband/Partner/Father* | 176 (67.4%) | 5089 (74.1%) | 0 (0%) | 0.72 (0.56, 0.95) | 0.017 |
| *Other relatives (sister/cousin, etc.)* | 50 (19.2%) | 1576 (22.9%) | 0 (0%) | 0.80 (0.58, 1.08) | 0.153 |
| *Friend* | 3 (1.1%) | 168 (2.4%) | 0 (0%) | 0.46 (0.11, 1.23) | 0.190 |
| *No substantial support at home* | 5 (1.9%) | 61 (0.9%) | 0 (0%) | 2.18 (0.76, 4.96) | 0.097 |
| **Prior history of poor milk production** | 5 (1.9%) | 206 (3%) | 8 (0.11%) | 0.63 (0.22, 1.39) | 0.314 |
| **Symptoms present at discharge** | 187 (71.6%) | 4103 (59.7%) | 0 (0%) | 1.70 (1.3, 2.25) | <0.001 |

**Abbreviations:** N = Number; SD = standard deviation; Q2 = Second Quartile (50th percentile); Q3 = Third Quartile (75th percentile); OR = Odds ratio; CI = Confidence interval; SpO2 = oxygen saturation; HIV = Human Immunodeficiency Virus; PNC = post-natal care; SES = Socio-economic index; WHO = World Health Organization

Sociodemographic factors also played an important role in their association with stillbirths. The strongest association was previous child death (OR: 7.11, 95% CI: 5.50–9.17, p < 0.001). The socioeconomic index score showed lower odds of stillbirth for each unit increase in the degree of socioeconomic index (OR: 0.81, 95% CI: 0.75–0.88, p < 0.001). Higher maternal education was also associated with lower rates of stillbirths, with mothers who had post-secondary education having lower odds of stillbirth (OR: 0.26, 95% CI: (0.14–0.53, p < 0.001), compared to mothers with less than a Primary three-level education.

Additionally, features related to geography and travel impacted the probability of stillbirth. Women who travelled more than an hour to reach the hospital (OR: 2.71, 95% CI: 1.95–3.75, p < 0.001) and those who relied on an ambulance (OR: 2.30, 95% CI: 1.54–3.38, p < 0.001) and arrival following a referral from a lower-level facility (OR: 3.44, 95% CI: 2.62–4.55, p < 0.001) had disproportionately higher stillbirths. Also, delays due to difficult terrain (OR: 4.48, 95% CI: 1.82–9.45, p < 0.001), general transport delays (OR: 2.12, 95% CI: 1.27–3.35, p = 0.002) and other unspecified delays (OR: 2.08, 95% CI: 1.01–3.81, p = 0.028) also increased risk.

### Factors associated with fresh and macerated stillbirth

Although the proportions of mothers experiencing macerated (43.7%) vs fresh (51%) stillbirths were similar, several maternal and delivery features were disproportionately represented. Mothers who lived with the baby’s father were less likely to have macerated stillbirths (OR: 0.37, 95% CI: 0.17–0.77; p = 0.009), as were those who had caesarean deliveries (OR: 0.24, 95% CI: 0.13–0.42, p < 0.001). Conversely, food insecurity during pregnancy (OR: 2.28, 95% CI: 1.20–4.43, p = 0.013), and higher education (p = 0.015) was associated with macerated stillbirths (Table 2).

**Table 2:**
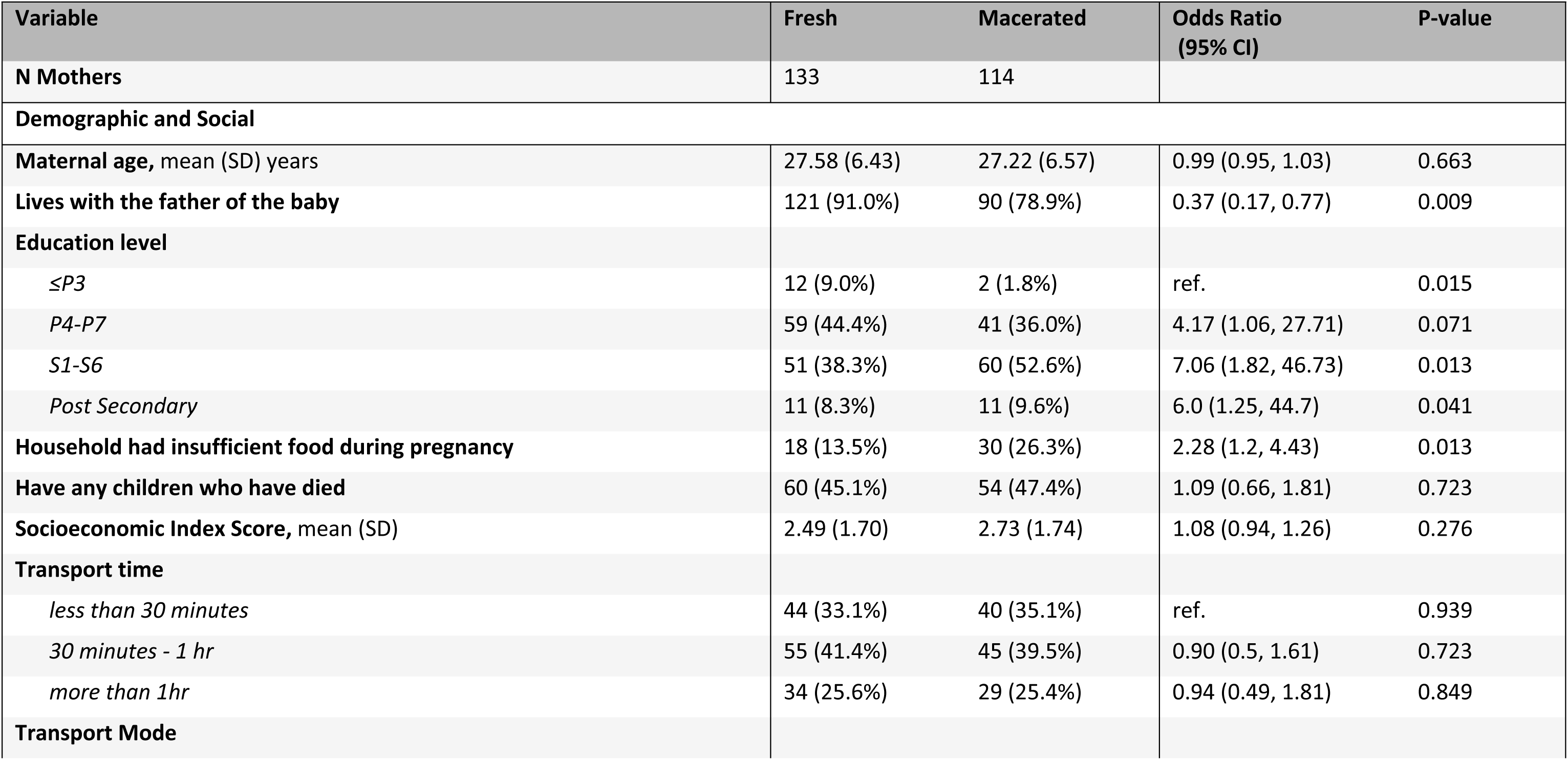
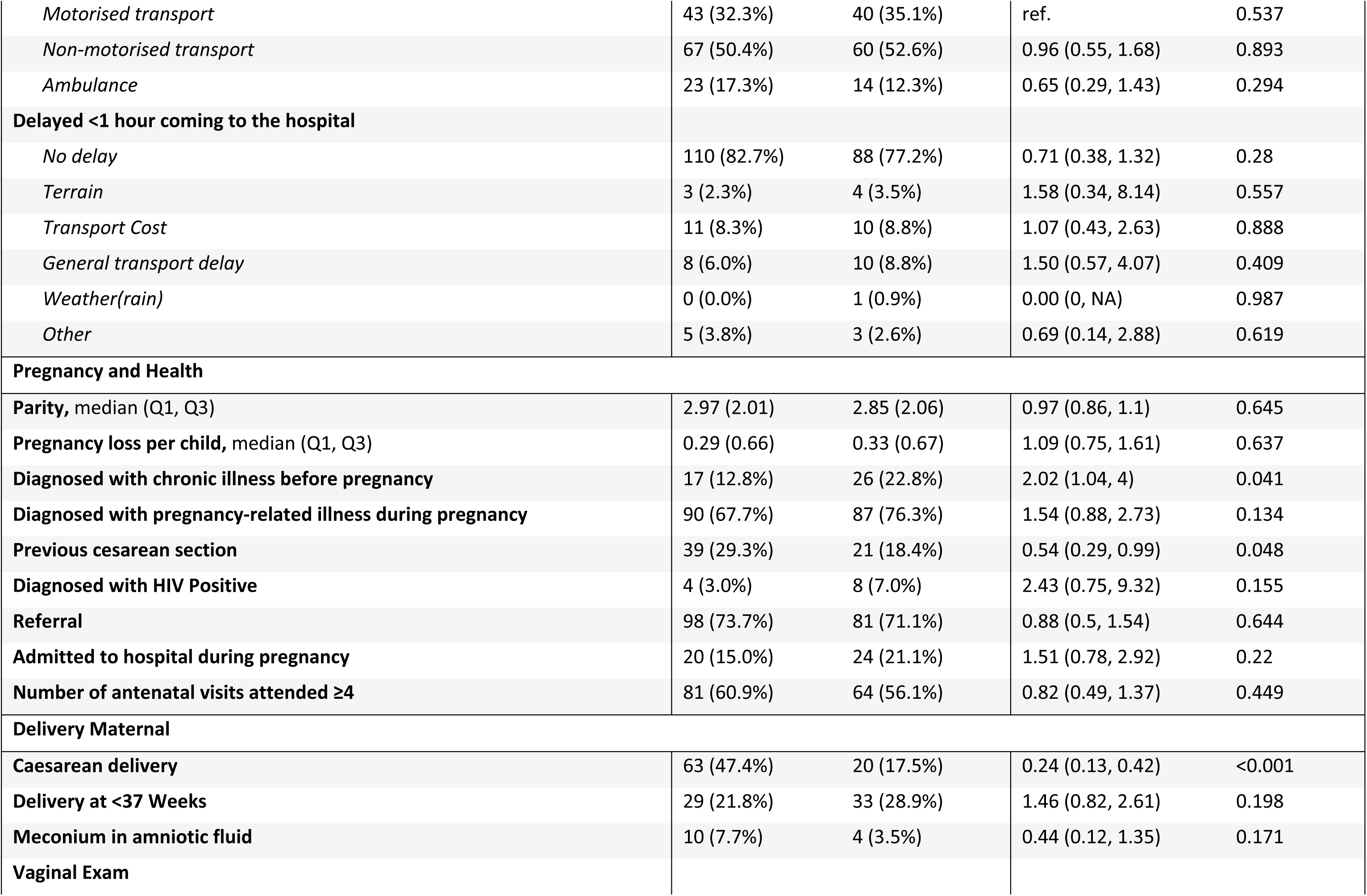
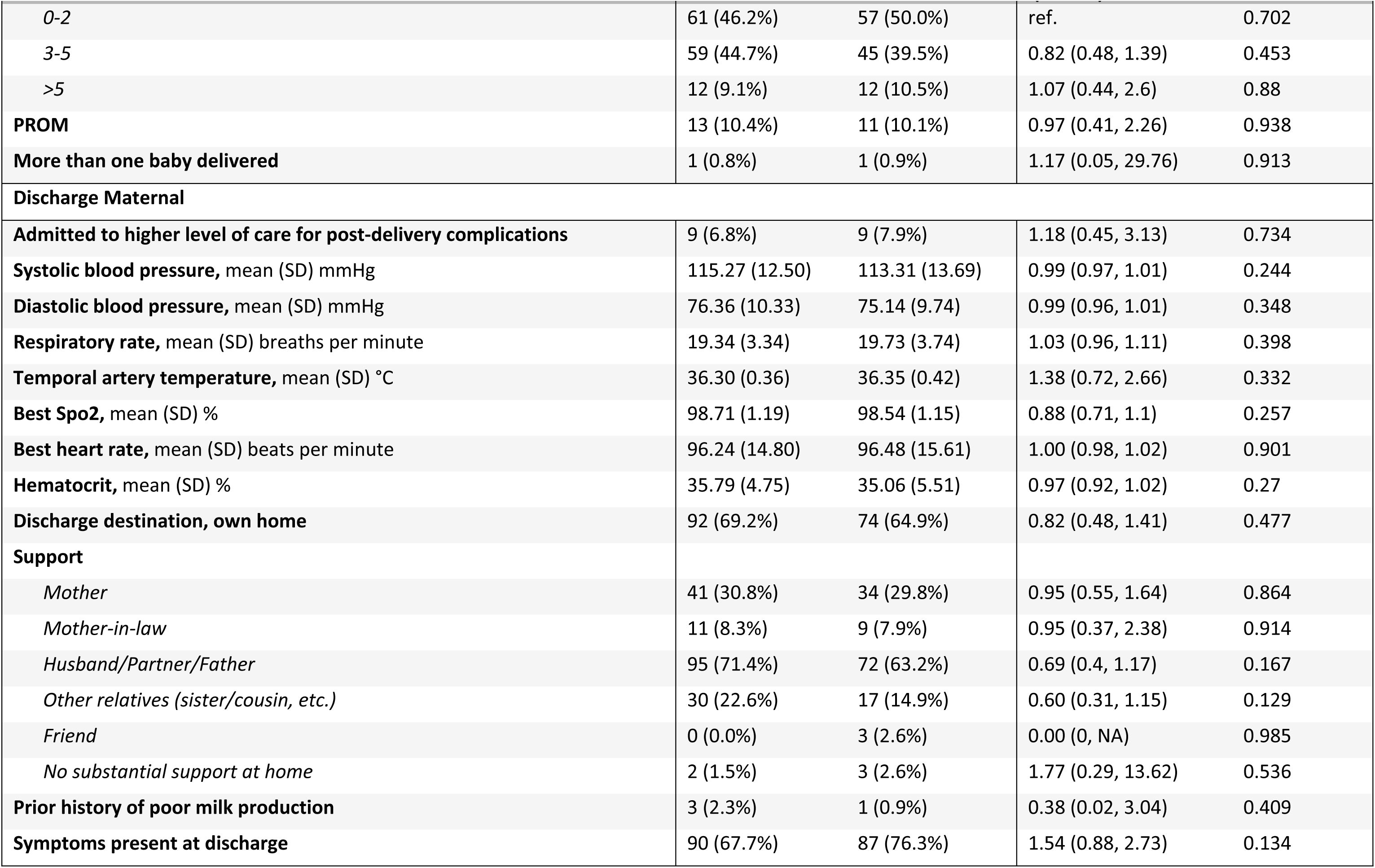
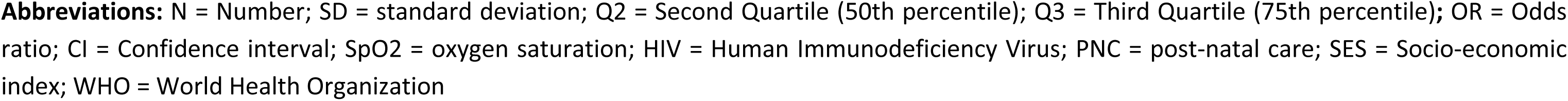
Maternal demographic, clinical, and delivery-related characteristics stratified by type of stillbirth (fresh vs macerated), with univariate odds ratios. The table presents maternal characteristics among women who experienced a stillbirth, stratified by fresh stillbirth and macerated stillbirth. Odds ratios (ORs) and 95% confidence intervals (CIs) were estimated using univariate logistic regression, with macerated stillbirth as the reference category. An OR >1 indicates higher odds of fresh stillbirth relative to macerated stillbirth. Continuous variables are summarized as mean (SD) or median (Q1, Q3), and categorical variables as n (%). The reference category for categorical variables is labeled “Ref.” For non-binary categorical variables, the global p-value is reported in the reference row.

### Clinical and maternal factors associated with in-hospital neonatal death

Similar features seen in the analysis of risk factors for stillbirths were seen in the analysis of risk factors for in-hospital newborn death. Previous child death (OR = 2.29, 95% CI: 1.42–3.56, p < 0.001) or pregnancy loss (OR = 1.28, 95% CI: 1.00–1.58, p = 0.029) as well as travel time exceeding one hour (OR = 2.09, 95% CI: 1.21–3.51, p = 0.007), and referral from another health facility (OR = 2.52, 95% CI: 1.72–3.76, p < 0.001) were associated with newborn mortality following a live delivery. Similarly, the mothers diagnosed with a pregnancy-related illness (OR = 1.53, 95% CI: 1.03–2.31, p = 0.037), premature rupture of membranes (PROM) (OR = 2.91, 95% CI: 1.50–5.16, p = 0.001) and multiple gestation (OR = 2.86, 95% CI: 1.33–5.44, p = 0.003) all were also associated with higher risk (Table 3).

## Discussion

This study reports a comprehensive analysis of the burden of, and risk factors for, stillbirth in a cohort of women presenting for delivery at two regional referral hospitals in Uganda. It also emphasizes the impact that stillbirths have on post-discharge trajectories. Among women experiencing a stillbirth, nearly 1 in 10 women required admission during the first 6 weeks following delivery. These results emphasize the urgent need for interventions that not only reduce the high rates of stillbirths but also improve postpartum care for women who experience stillbirths.

The present analysis affirms prior research that has shown that social and geographic vulnerability, alongside clinical risk factors, are important features that are associated with stillbirths (1). Lower socioeconomic index scores, delayed access to health facilities due to terrain features (lakes, floods, poor roads, etc.), and timely access to transportation were all important risk factors for stillbirth (13). A history of previous child death was the strongest predictor of stillbirth in the current cohort. Women who had such a history had over seven times the odds of stillbirth compared to those with no prior history of a loss. These results reflect the multifactorial nature of stillbirths and reinforce the call for strengthening the health system to tackle modifiable factors and improve outcomes.

In this analysis, we found that just over half of stillbirths were classified as fresh, suggesting fetal demise during labor or delivery. This is consistent with prior research in resource-limited settings (14) and points to critical gaps in intrapartum care, particularly fetal monitoring and management of labor complications (15,16). In contrast to the fresh stillbirths, the macerated stillbirths suggest antepartum causes, and our results point to these being tied to socioeconomic and clinical vulnerabilities, demonstrating the clear need for the strengthening of antenatal care services. Overall, stillbirths in general were associated with meconium-stained amniotic fluid, preterm delivery, and transportation delays, suggesting delays in recognition and response to fetal distress. These results largely align with studies in sub-Saharan Africa that link intrapartum stillbirths to inadequate labor monitoring, obstructed labor, and limited access to emergency obstetric care (17).

This analysis also identifies a range of maternal, environmental, and clinical factors that significantly influence in-hospital neonatal mortality, many of which reflect systemic barriers to accessing timely and quality care. Environmental delays, especially those caused by adverse weather conditions and difficult terrain, are key obstacles to reaching a health facility (18,19). Clinically, preterm birth remains one of the strongest predictors of neonatal death, consistent with known risks such as respiratory distress, immaturity, and increased infection vulnerability among premature babies (20,21). A history of prior child death also raises the risk of neonatal mortality, indicating persistent maternal or fetal risk factors like chronic illness or limited antenatal care that can affect subsequent pregnancies (22,23). From a health system view, referrals from lower-level facilities are linked to poorer neonatal outcomes, reflecting delays in triage, transportation, and emergency coordination. Given that caesareans are often life-saving, their association with neonatal mortality here likely indicates emergency cases and complications related to the procedure (24,25).

Postpartum readmission was significantly more common among women who experienced stillbirth compared to those with live births, highlighting a potential area for targeted postpartum support. Women readmitted after stillbirth were more likely to report antenatal morbidity and socioeconomic hardship, suggesting that the cumulative burden of medical and social vulnerabilities may increase the risk of complications following discharge. These findings are consistent with population-based studies that have demonstrated higher rates of postpartum readmission among women with stillbirth, largely driven by unresolved physical complications, such as infection and hemorrhage, as well as psychological distress (26). Moreover, prior research has emphasized that bereaved mothers often experience limited social support and face barriers to accessing appropriate postpartum follow-up care, compounding their risk of readmission (27). These results underscore the importance of integrating mental health services and individualized medical follow-up into the postpartum care pathway for women who experience stillbirth, particularly those with underlying antenatal complications or socioeconomic disadvantage.

This study has several limitations. First, the observational design restricts our ability to determine causality between the identified risk factors and stillbirth or postpartum readmission. Second, data were collected from two major regional referral hospitals that serve both urban and rural populations. Although we believe these hospitals to be broadly representative of the Ugandan population, the results may lack generalizability to lower-tier health facilities or private healthcare facilities where barriers to care and outcomes may differ. Lastly, self-reported measures of socioeconomic status and care-seeking behavior can be influenced by recall and social desirability biases, especially among women who have experienced stillbirth, potentially affecting the accuracy of these responses.

In conclusion, this study reports important insights into the multifactorial contributors to stillbirth and postpartum readmission in a low-resource setting. Our findings underscore the critical role of both clinical and social determinants, including previous child loss, preterm delivery, antenatal morbidity, and care-seeking delays in shaping maternal and perinatal outcomes. The distinct patterns observed between intrapartum and antepartum stillbirths highlight the need for context-specific strategies to strengthen intrapartum care and address upstream social vulnerabilities. Moreover, the significantly higher postpartum readmission rates among women who experienced stillbirth point to unmet health needs and potential gaps in post-discharge care. These results call for integrated interventions that prioritize early risk identification, improve access to emergency obstetric care, and extend support beyond hospital discharge, especially for bereaved mothers. Investing in such targeted efforts will be essential to reducing preventable stillbirths and improving long-term maternal health outcomes. These findings also collectively emphasize the intricate factors leading to neonatal mortality and the pressing need to improve maternal and newborn care across the entire continuum, from antenatal monitoring to facility-based delivery and postnatal care.

## Data Availability

Study materials, such as de-identified data, a data dictionary, and analysis code, can be made available upon reasonable request to the corresponding author or through the Paediatric Sepsis CoLab. The University of British Columbia Dataverse Collection: Paediatric Sepsis CoLab. Smart Discharges Dataverse. Borealis. 2022. https://borealisdata.ca/dataverse/smart_discharge. Additional data cannot be publicly shared due to ethical restrictions involving participant confidentiality.

## Acknowledgments

We express our gratitude to the study participants, healthcare providers, and data collection teams from the two regional referral hospitals (MRRH, JRRH). Additionally, we acknowledge the support of the Lacuna Fund, which facilitated this secondary data analysis as part of a broader initiative to enhance capacity in data science for health research in low-resource settings. We also appreciate our institutional collaborators for their technical input throughout the study.

## Contributors

OT was responsible for statistical analysis and writing the first draft the manuscript; MOW and VN assisted with the development, writing, and editing of the manuscript as well as coordinating author contributions. VN and MOW supervised data analyses. VN, OT, AN, MOW, JN, CK, AC-D interpreted the data, and OT, MOW, VN, CK, YP, AN, JN, AC-D critically reviewed the manuscript.

## Supporting information

**S1 Table.** Factors for neonatal in-hospital death.

**S2 Table.** Maternal readmission after stillbirth.

**S1 File.** STROBE checklist for cohort studies.

